# Health inequalities in SARS-CoV-2 infection during the second wave in England: REACT-1 study

**DOI:** 10.1101/2023.08.01.23293491

**Authors:** Haowei Wang, Kylie E. C. Ainslie, Caroline E. Walters, Oliver Eales, David Haw, Christina Atchison, Claudio Fronterre, Peter J. Diggle, Deborah Ashby, Graham Cooke, Wendy Barclay, Helen Ward, Ara Darzi, Christl A. Donnelly, Steven Riley, Paul Elliott

## Abstract

**Background:** The rapid spread of SARS-CoV-2 infection caused high levels of hospitalisation and deaths in late 2020 and early 2021 during the second wave in England. Severe disease during this period was associated with marked health inequalities across ethnic and sociodemographic subgroups.

**Methods:** We analysed risk factors for test-positivity for SARS-CoV-2, based on self-administered throat and nose swabs in the community during rounds 5 to 10 of the REal-time Assessment of Community Transmission-1 (REACT-1) study between 18 September 2020 and 30 March 2021.

**Results:** Compared to white ethnicity, people of Asian and black ethnicity had a higher risk of infection during rounds 5 to 10, with odds of 1.46 (1.27, 1.69) and 1.35 (1.11, 1.64) respectively. Among ethnic subgroups, the highest and the second-highest odds were found in Bangladeshi and Pakistan participants at 3.29 (2.23, 4.86) and 2.15 (1.73, 2.68) respectively when compared to British whites. People in larger (compared to smaller) households had higher odds of infection. Health care workers with direct patient contact and care home workers showed higher odds of infection compared to other essential/key workers. Additionally, the odds of infection among participants in public-facing activities or settings were greater than among those not working in those activities or settings.

**Interpretation:** Planning for future severe waves of respiratory pathogens should include policies to reduce inequality in risk of infection by ethnicity, household size, and occupational activity.

## Introduction

The COVID-19 pandemic has caused a substantial increase in hospitalisations and mortality globally since its emergence in China in late December 2019 and subsequent spread.[1] Populations continue to experience multiple waves of infection, with substantial reactive changes in social distancing still occurring in some countries either spontaneously or as a result of explicit government regulations.[2,3] Social distancing measures led to increased inequalities before the pandemic hit and interacted with pre-existing structural inequalities such as sex, age, region and ethnicity. [4,5] Investigating how inequalities influence the risk of getting infected is crucial for policymakers in understanding the complex impacts of the pandemic and shaping their response to it.

In order to enable isolation and quarantine policies, routine testing programmes were implemented nationwide, which mainly targeted individuals who reported symptoms.[6] While targeted testing provides valuable surveillance, sub-clinical cases that are likely to contribute substantially to transmission are not captured. Furthermore, the uptake of testing may be limited by test capability and geographic, demographic and social factors, and thus not be fully representative even among those eligible for tests. Therefore, tracking the characteristics and underlying inequalities of individuals at risk of SARS-CoV-2 infections at the population level can provide additional evidence for policymaking.[7] In the UK, large representative population-based studies were conducted in the UK to better understand the underlying trend of the epidemic and risk factors.[8–11]

There is considerable evidence across many geographies that minority ethnic groups were at increased per-person risk of severe disease and death from COVID-19. However, there remains considerable uncertainty as to what extent this was driven by differential exposure,[12–14] susceptibility,[15–17] infection,[15,18,19] access to testing,[20,21] or differential access to health care.[21–23] Reducing these disparities during future similar emergencies will only be possible if we understand why they arose during this pandemic.

The REal time Assessment of Community Transmission-1 (REACT-1) study was designed to be representative of the population of England as a whole.[24] The REACT-1 study tracked the spread of the virus through a total of 19 cross-sectional surveys to collect self-administered throat and nose swabs which were tested for the presence of SARS-CoV-2 by reverse transcriptase polymerase chain reaction (rt-PCR).[24] The first round of the study began on 1 May 2020, and each new round thereafter was conducted at approximately monthly intervals, with a two-week break between the end of the previous round and the beginning of the next round. In each round up to round 12, participants were sampled randomly with a similar achieved sample size from 315 lower-tier local authorities (LTLAs). Participants were asked to take part in the study regardless of whether they were symptomatic or not.

Here, we present findings on risk factors and quantify the differences in risk of SARS-CoV-2 infection for rounds 5 to 10 between mid-September 2020 and the end of March 2021. Results on risk factors from the previous four rounds between May and the beginning of September 2020 have been published previously.[25]

## Methods

### Data collection

The detailed study protocol of the REACT-1 study has been reported elsewhere.[24] In brief, we randomly selected individuals aged 5 years and above from the National Health Service (NHS) patient list to send personalised invitations, balanced across 315 lower-tier local authority areas (LTLAs). Participants received test kits and instructions delivered by post and conducted self-administered throat and nose swabs (parent/guardian administered at ages 5 to 12 years). In addition to the information registered on the NHS list (name, age, sex, and address), participants were requested to complete an online or telephone questionnaire to provide other detailed information, e.g. employment type, ethnicity, household size, whether they were in contact with a confirmed or suspected COVID-19 case and symptom status.

### Socio-demographic groups

Study questionnaires are available on the REACT 1 Study Materials webpage on the Imperial College London website.[26]

In order to investigate prevalence by subgroups, we converted continuous variables such as age into categories and regrouped some categorical variables into fewer categories by combining similar categories. We divided age into eight age groups as follows: 5 to 12, 13 to 17, 18 to 24, 25 to 34, 35 to 44, 45 to 54, 55 to 64 years, and 65 years or older. Nineteen ethnic groups were re-grouped into five broad categories, white, Asian, black, mixed ethnic group, and other ethnic groups for some analyses. Participants were asked to report the number of children or young people between the ages of 0 and 17 and adults aged over 18 years who currently live in or spend part of the week in the household of the respondent. We obtained the total household size by adding these numbers. We then created a new categorical variable for household size with 6 categories (1 person, 2 people, 3 people, 4 people, 5 people and 6 or more people), based on the number of people in each household. For the variable on COVID-19 case contact, participants answered whether they were in contact with a confirmed/tested or suspected COVID-19 case. We did not further re-categorize this variable.

We asked participants if they worked as health care workers or care home workers or other key workers. If they did not work as a key worker, they were asked if they were engaged in one or more public-facing activities or settings (11 different activities for rounds 5 to 8, 12 activities for rounds 9 and 10). In addition, participants were asked about their employment status (employed in full-time jobs, employed in part-time jobs, self-employed, unemployed, retired from work). We combined employment and key worker answers into a new single variable which we called key worker status that could take one of the following four values: 1) Health care or care home worker, 2) Other essential/key workers, 3) Other workers, and 4) Not full-time, part-time, or self-employed.

We also regrouped symptom status during the month before the test. Participants were classified as ‘Classic COVID-19 symptoms’ if they had experienced one or more of the following symptoms in the past month: loss or change of a sense of smell or taste, persistent cough, or fever. Other symptomatic people who did not show classic COVID-19 symptoms in the past month were classified as ‘Other symptoms’. Participants who did not feel unwell and did not present any of the listed symptoms were classified as ‘No symptoms’.

### Statistical analysis

We estimated the unweighted prevalence as the proportion of the total samples that were positive. To correct for non-response bias and unequal selection probabilities that occurred in the sampling process, we used Random Iterative Method (RIM) weighting[27] to adjust unweighted prevalence with pre-calculated survey weights and obtain prevalence estimates representative of the population in England. The method, and the process of applying it to REACT-1 data, have been described elsewhere.[25] RIM weights were based on age, sex; LTLA response; ethnic group; and IMD (Index of Multiple Deprivation[28]) deciles.

We fitted multivariable logistic regression models to investigate whether particular subgroups were more likely to test positive for SARS-CoV-2. The binary outcome variable was swab-positivity (yes or no), while age, sex, region, broad categories of employment, broad categories of ethnicity, household size and deprivation were included as explanatory variables. The odds ratio (OR) was reported along with corresponding 95% confidence intervals. The associations of swab-positivity with detailed categories of occupation and ethnicity subgroups were also quantified using logistic regression models: for detailed occupation, the model was adjusted for age, sex, region, broad categories of ethnicity, and deprivation; and for ethnicity subgroups, the model was adjusted for age, sex, region, broad categories of employment, and deprivation.

To assess the overall risk for subgroups across rounds 5 to 10, we pooled log-odds ratios from each individual round using the inverse-variance method.[29]

## Results

### Levels of swab-positivity during the second wave in England

England experienced a large second wave of SARS-CoV-2 infection between September 2020 and March 2021, with rising overall swab positivity from September 2020 to January 2021, followed by a decline to March 2021 (Figure 1, Table S1). Between 18 September 2020 and 30 March 2021, 7,052 samples were positive overall from 977,245 swabs.

**Figure 1.**
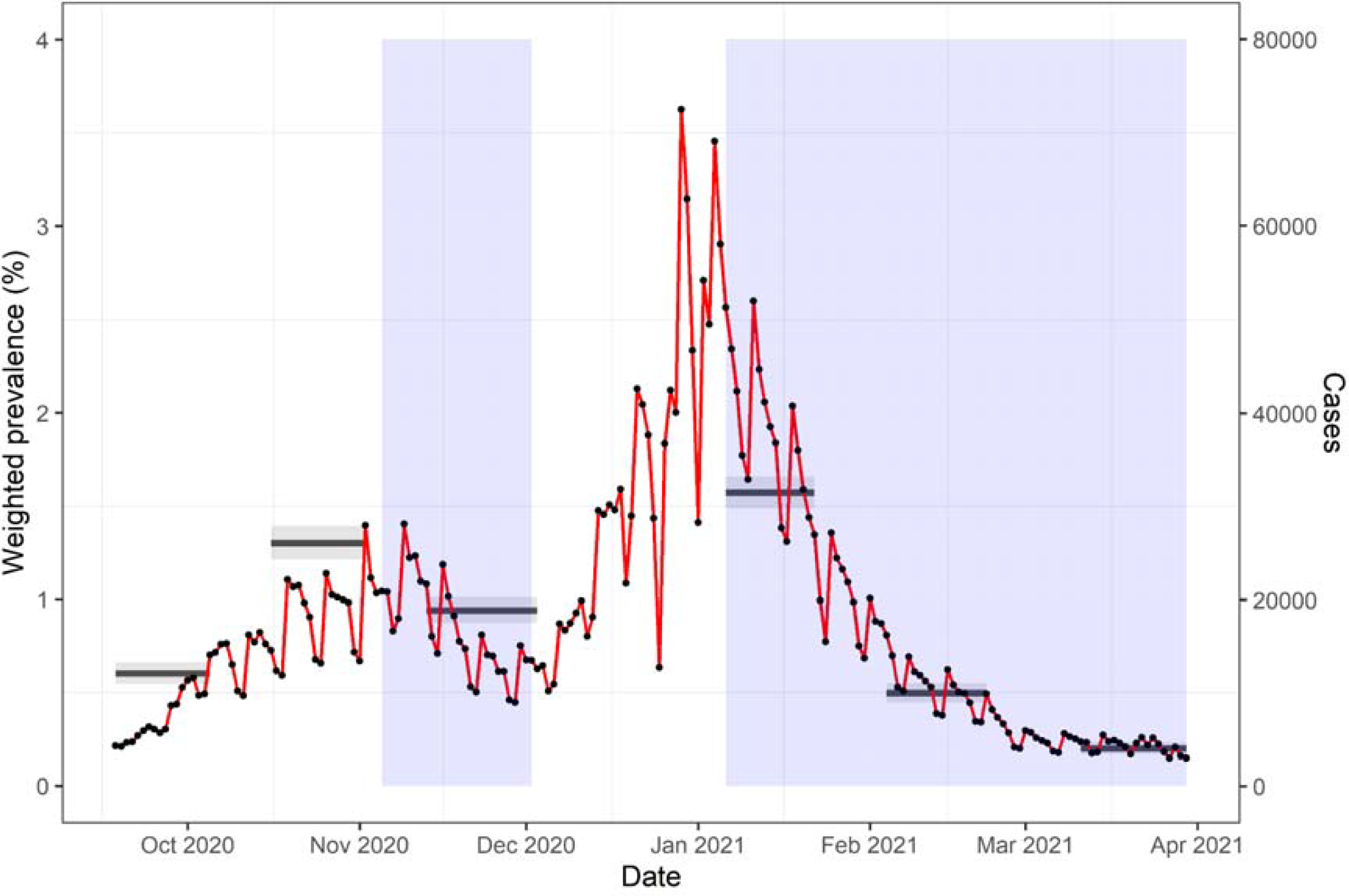
Comparison of overall weighted prevalence as measured by REACT-1 to surveillance case data (Public Health England, PHE). Overall weighted prevalence for rounds 5 to 10 of the REACT-1 study is in black solid lines (left-hand side y-axis) with a 95% confidence interval (grey shaded boxes). The temporal dynamic of surveillance data is shown by black points and the red solid line (right-hand side y-axis). The right y-axis is scaled by 20,000 per case. Blue-shaded areas indicate the time period of the second and the third lockdowns in England.

### Ethnicity

Self-identified broad classes of ethnicity were a substantial risk factor for testing positive during this period of the epidemic in England and remained important even after adjustment for household size and level of local neighbourhood deprivation. Broad classes of ethnicity were defined by participants self-identifying as one of the white, Asian, black, mixed and other ethnic groups. The Asian ethnic group showed the highest weighted prevalence consistently during the second wave, while the white ethnic group was at the lowest level (Figure 2A, Table S2). Similar patterns of higher infection prevalence in black and Asian ethnic groups were seen in England in subsequent REACT-1 rounds.[30–38] Across rounds 5 to 10, the odds of being swab-positive were 1.45 (1.27, 1.66) and 1.34 (1.11, 1.63) times greater for people of Asian and black ethnicity respectively, compared to white ethnicity, whereas the odds were 0.93 (0.77, 1.11) times lower for people of mixed ethnic group than white ethnic group (Figure 2A, Table S3).

**Figure 2.**
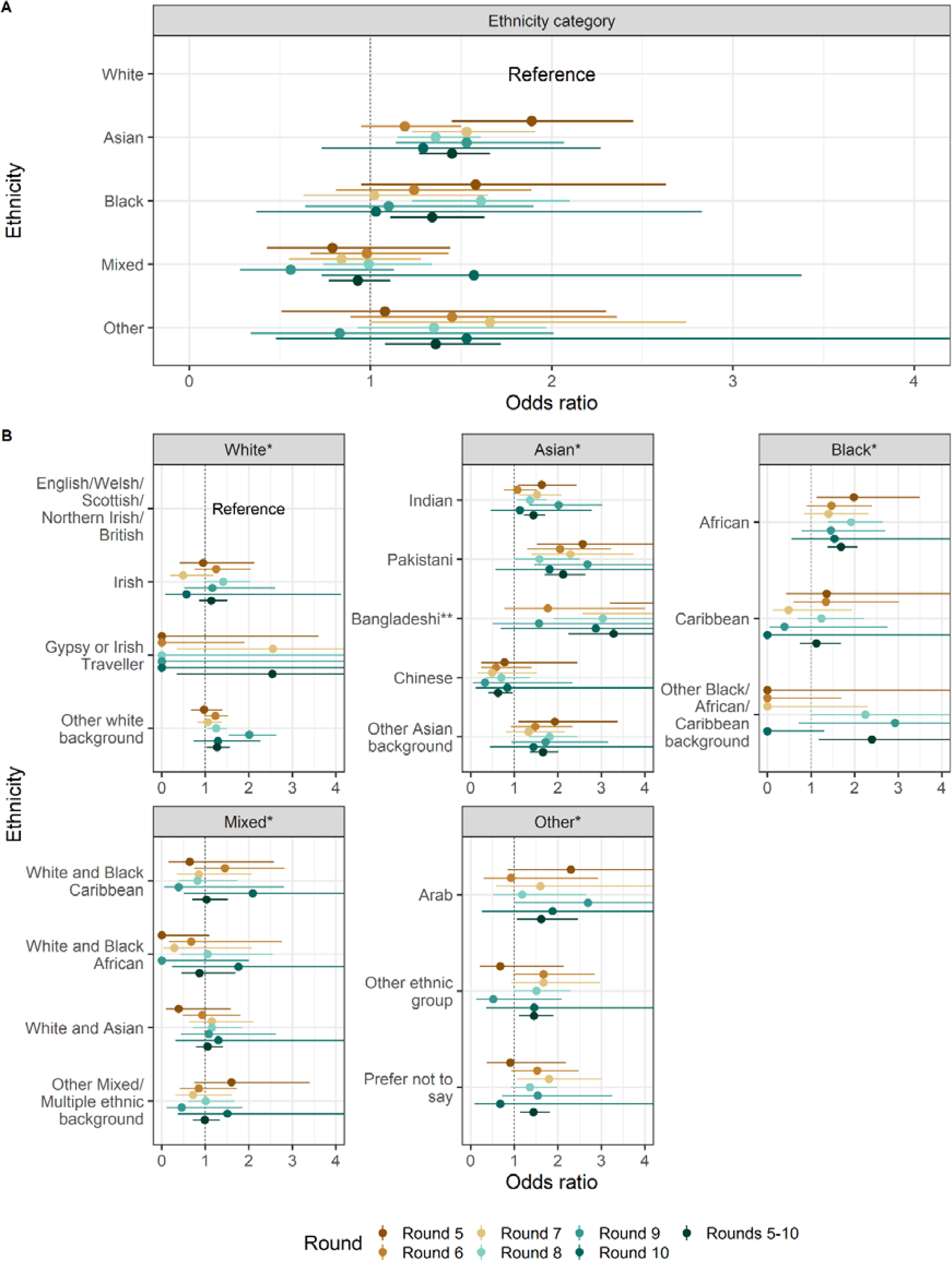
Forest plot of ethnicity categories and detailed subgroups of ethnicity for rounds 5 to 10 and an average across rounds weighted inverse to variance (rounds 5 to 10). **A:** Forest plot of ethnicity categories, where the reference group is white ethnicity. **B:** Forest plot of detailed subgroups of ethnicity, the reference group is those identifying as English, Welsh, Scottish, Northern Irish or British. Individual round odds ratios are mutually adjusted for sex, age group, region, key worker status, and deprivation index. * For detailed subgroups of ethnicity, they are included in one model where the reference group is those identifying as English, Welsh, Scottish, Northern Irish or British. ** The odds ratio for Bangladeshi ethnicity in round 5 was 5.75 (3.29,10.07)

Participants were also asked to self-identify as a member of an ethnic subgroup. Some self-identified ethnic subgroups had a higher prevalence of SARS-CoV-2 infection in some rounds and lower in others (Table S4). These temporal differences were substantial and likely reflected underlying epidemic dynamics,[39] rather than chance variation. For example, from rounds 5 to 8, the prevalence of swab-positivity in Bangladeshi participants was very high, reaching 6.12% (3.78%, 9.29%) in round 8. By comparison, the prevalence among white ethnic subgroups was less than 2%, with the highest value found in white Irish participants at 1.93% (1.30%, 2.74%). During the same period, black ethnic subgroups also showed a higher prevalence than white ethnic subgroups.

Excess risk of infection was also seen in these ethnic subgroups compared with the British white populations, even after accounting for the differences in age, sex, region, employment status, household size and level of local neighbourhood deprivation. Among subgroups, across rounds 5 to 10, the highest and the second-highest odds were found in Bangladeshi and Pakistani ethnic subgroups at 3.28 (2.24, 4.8) and 2.12 (1.7, 2.64), respectively, compared with British white participants, which contributed to the high odds in Asians (Figure 2B, Table S5).

### Household size and deprivation

Participants in larger households consistently had higher rates of infection than in smaller households. Prevalence increased monotonically with household size, and households with six or more people consistently had the highest prevalence in rounds 5 through 10. These results were maintained when estimating odds ratios adjusted for age and sex, where participants in households of 3 to 5 people and 6 or more people had overall increased odds of 1.30 (1.18, 1.43) and 1.32 (1.21, 1.44), respectively, compared with participants in households of size 1 or 2. This trend was accentuated by a model adjusted for region, key worker status, ethnicity and deprivation in addition to age and sex, with odds ratios of 1.92 (1.64, 2.24) and 1.80 (1.60, 2.03) respectively. (Figure S1.B, Table S3).

In rounds 5 to 10, participants in the most deprived areas consistently had a higher risk of infection than those in the least deprived areas, with an overall increased odds of 1.69 (1.51, 1.89) adjusted for age and sex. The deprivation difference remained after adjusting for age, sex, region, broad categories of employment type, broad categories of ethnicity, and household size, with people in the most deprived quintile having an increased odds of 1.42 (1.23, 1.63) compared to the least deprived (Figure 3, Table S3).

**Figure 3.**
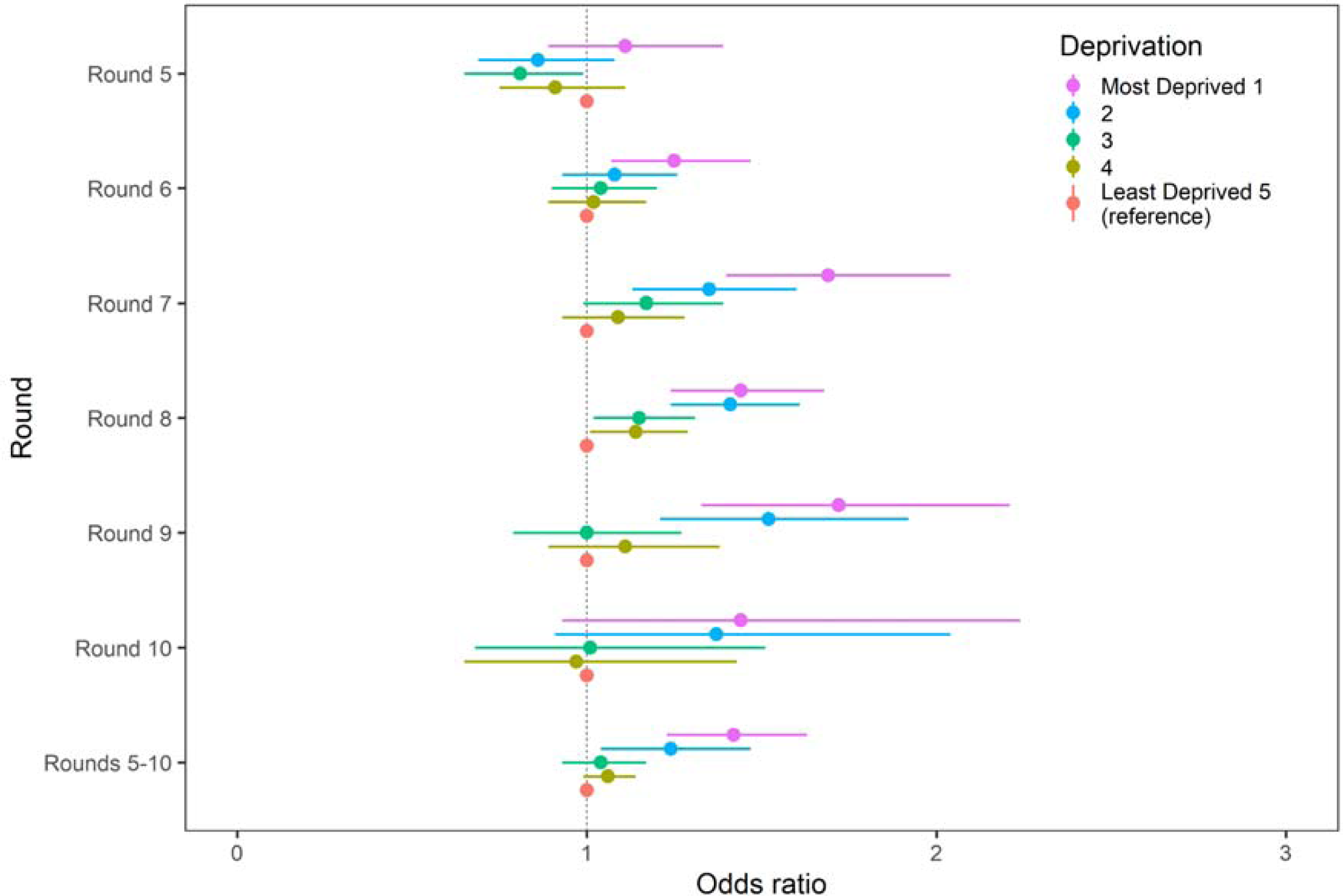
Forest plot of deprivation for rounds 5 to 10 and an average across rounds weighted inverse to variance (rounds 5 to 10). The reference group is those living in the least deprived areas (Least Deprived 5). Individual round odds ratios are mutually adjusted for sex, key worker status, broad categories of ethnicity, household size and deprivation index.

### Occupation

Occupation was also a risk factor for swab-positivity. Across rounds 5 to 10, compared to non-key workers, the odds of infections in health care and care home workers was 1.32 (1.13, 1.54) (Table S3). Among key workers, health care workers with direct patient contact had increased odds of 1.18 (1.04, 1.33), while the odds for health care workers with no patient contact was 0.77 (0.6, 0.99), compared with other essential/key workers. The odds of swab-positivity were increased among care home workers, both those with direct contact with clients at 1.35 (1.01, 1.81) and those without contact with clients at 2.44 (1.57, 3.79) compared to other essential/key workers (Figure 4A, Table S6 - 7).

**Figure 4.**
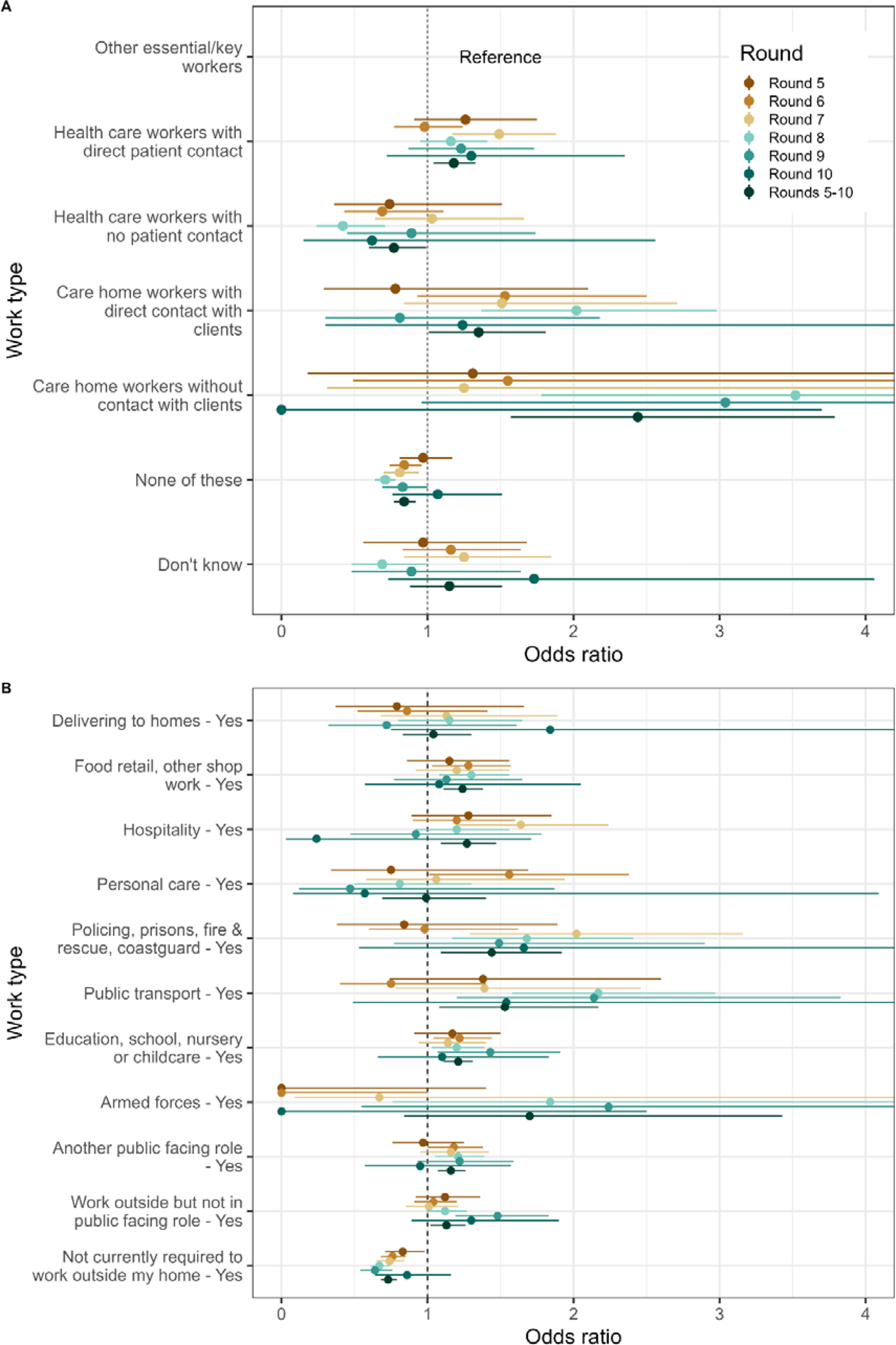
Forest plot of occupations for rounds 5 to 10 and an average across rounds weighted inverse to variance (rounds 5 to 10). **A:** Reference group is other essential/key workers. Note that the bottom four categories are combined in Figure 4B as HCW/CHW, and combined with full-time and part-time status in the variable named ‘key worker status’. **B:** Only people who did not work in health or social care (top four boxes, Figure 6. A) were asked whether their work involved these activities or settings. Each work activity or setting has a different reference group - those who answered that they did not work with that activity or in that setting. For example, the reference group for those that answered that they delivered to homes was the group who answered that they did not deliver to homes. Individual round odds ratios are mutually adjusted for sex, age group, region, broad category of ethnicity, deprivation index and the specific work activity or setting represented.

We also asked non-key workers about specific activities that they may or may not have had to undertake related to their work. For example, compared to those not occupationally exposed to the activity, participants whose work activity included: food retail or other shop work had increased odds of 1.24 (1.11, 1.38); hospitality had odds of 1.27 (1.09, 1.47); public transport had odds of 1.53 (1.08, 2.17); and participants working in education, school, nursery or childcare had odds of 1.21 (1.11, 1.31) (Figure 4B, Table S7). The odds of infections for participants who were not required to work outside their homes were lower at 0.73 (0.68, 0.79) compared with those who were required to work outside their homes.

### Sensitivity analysis for vaccination status

As the rollout of the vaccination programme in England began on 8 December 2020,[40] we included vaccination-related questions in the questionnaire from round 8 (January 2021) and asked participants for consent to longer-term follow-up through linkage to their NHS records. Due to the low number of people vaccinated in round 8, we only linked to NHS vaccination records during rounds 9 and 10 with a match to 114, 747 (69%) participants in round 9 and 98,983 (70%) participants in round 10.

We compared the results from the regression models reported above adjusted for sex, age, region, key worker status, ethnicity, household size and neighbourhood deprivation, with similar models also adjusted for vaccination status. There were no marked differences between these two sets of models (Table S8). For example, the odds of SARS-CoV-2 infection remained higher among health care and care home workers at 1.87 (1.03, 3.38) after adjustment for vaccination status. As expected, we also found participants who received one or two doses of vaccination had lower odds of infection than unvaccinated participants, with odds ratios (ORs) of 0.63 (0.45, 0.86) and 0.61 (0.23, 1.59) respectively.[32]

## Discussion

In this large community-based study of SARS-CoV-2 infection in England, we found during the period September 2020 to March 2021 that minority ethnic subgroups, larger household sizes, increasing neighbourhood deprivation, and key worker status were independently associated with increased odds of testing positive for SARS-CoV-2. During that time, few people were vaccinated, and the perceived risk of infection was high. Our results suggest that substantial inequalities arose because of differential risk of infection rather than access to testing and healthcare.

Our findings build on prior descriptions of antigenic exposure to SARS-CoV-2 in a similar study in England during the period immediately prior to September 2020,[41] which suggested that higher hospitalisation and deaths in ethnic minority subgroups were because of their higher infection risk, not because they were intrinsically more vulnerable. Therefore, the results presented here imply that health inequality driven by a higher risk of infection persisted in England from September 2020 through to the following winter, during which there was high excess mortality. [42]

Factors driving increased infection in ethnic minorities are not immediately obvious: for example, one study of social mixing patterns during 2020 [43] found reduced rates of mixing in some ethnic groups, rather than increased rates which would provide a mechanism for increased rates of infection. However, this may have been driven by response bias during a period when many people were at home with more time to enrol in surveys compared with those going out to work.[44] In contrast, a US study analysed mobility data as a proxy for mixing at the level of a county, using variation in the ethnic diversity in each county to infer relative levels of mixing for different ethnic groups.[45] They found that African Americans had the lowest reduction in mobility and the highest rates of severe disease compared with white and Asian American populations.

These results represent substantial evidence that variation in infection drives variation in disease among ethnic groups. While, other studies have speculated that other factors explain differences in patterns of disease, such as genetic susceptibility,[46–48] treatment access, and comorbidities,[49] such as obesity,[50] diabetes,[51] and hypertension,[52] few studies are representative or measure infection, making it difficult to identify the root causes of increased disease accurately. Further studies are warranted to better characterise the contribution of these different factors to health inequality during the current post-pandemic phase of SARS-CoV-2 circulation.

Our study has limitations. First, we aimed to obtain representative samples for the population of England. Although we were able to correct for variations in response and estimate weighted prevalence for the population of England and specific subgroups, the number of people taking part in some subgroups was relatively low; this resulted in higher uncertainty in the estimates (with wider 95% confidence intervals). For example, we chose to present weighted prevalence only if the number of positives in a category was 10 or more; among people of ethnic minorities during some periods, the number of positives was too small to obtain robust estimates for weighted prevalence using classic survey-weighting methods. As a result, some important differences between subgroups were not fully captured. Although other analytical approaches, such as Bayesian hierarchical regression, may be better able to infer patterns where the sample size is small,[53] they would also require additional assumptions about the relationship between hierarchies, which we felt were not justified.

Also, the effects of risk factors might vary over time, for example, due to changes in contact or behavioural patterns among subgroups. Thus, students returning to universities from home after the holiday will increase contacts while travelling and on arrival at university. In addition, as the vaccine programme in England prioritised certain subgroups of the populations, they might have acquired sufficient immunity to reduce the risk of infections [54] in ways not captured by our sensitivity analyses. Although we investigated odds ratios for risk factors by rounds, we did not investigate within-round temporal trends in the relationship between risk factors and swab positivity.

Furthermore, response rates were lower for minority ethnic groups. In theory, this could have biased these results. However, we would expect such bias to have decreased rates of infection in these groups, rather than increased them, because people who take part in surveys tend to have lower rates of exposure.[55] Further, being present at home during lockdown periods made participation in our study easier, and less likely to have been in contact with other people.[56,57]

In conclusion, our findings show substantial differences in the risk of SARS-CoV-2 infections among minority ethnic groups and other subgroups of the population in England during a period when there was a substantial risk of disease and death from COVID-19. Planning for future severe waves of respiratory pathogens should include policies to reduce inequalities driven by ethnicity, household size, and occupational activity. [58]

## Contributors

SR and PE are joint corresponding authors. SR, CAD and PE conceived the study and the analytical plan. HW, KECA, CEW, OE, and SR performed the statistical analyses. HW, KEA, CEW and OE curated the data. HW, KECA, CEW, OE, CA, CF, DA, WB, GT, GC, HW and AD provided insights into the study design and results interpretation. AD and SR obtained funding. All authors revised the manuscript for important intellectual content and approved the submission of the manuscript. SR had full access to the data and took responsibility for the integrity of the data and the accuracy of the data analysis and for the decision to submit for publication.

## Data availability statement

Access to REACT-1 data are restricted because of ethical and security considerations. Summary statistics and descriptive tables from the current REACT-1 study are available in the appendix and online. Data from each round of the REACT-1 programme are summarised online. Additional summary statistics and results from the REACT-1 programme are also available online. REACT-1 study materials are also available for each round online at https://www.imperial.ac.uk/medicine/research-and-impact/groups/react-study/react-1-study-materials/.

## Ethics, consent and public involvement

The REACT programme studies obtained research ethics approval from the South Central-Berkshire B Research Ethics Committee (IRAS ID: 283787). Participants provide informed consent when they register for the studies, and all data are handled securely in accordance with a detailed privacy statement. A REACT Study Public Advisory Panel meets fortnightly to provide ongoing input into the research design, delivery and dissemination.

## Data Availability

https://github.com/mrc-ide/reactidd/tree/master/inst/extdata

https://www.imperial.ac.uk/medicine/research-and-impact/groups/react-study/real-time-assessment-of-community-transmission-findings/

https://www.imperial.ac.uk/medicine/research-and-impact/groups/react-study/react-1-studymaterials/

## Acknowledgements

SR, CAD acknowledge support: MRC Centre for Global Infectious Disease Analysis, National Institute for Health Research (NIHR) Health Protection Research Unit (HPRU), Wellcome Trust (200861/Z/16/Z, 200187/Z/15/Z), and Centres for Disease Control and Prevention (US, U01CK0005-01-02). GC is supported by an NIHR Professorship. HW acknowledges support from an NIHR Senior Investigator Award and the Wellcome Trust (205456/Z/16/Z). PE is Director of the MRC Centre for Environment and Health (MR/L01341X/1, MR/S019669/1). PE acknowledges support from Health Data Research UK (HDR UK); the NIHR Imperial Biomedical Research Centre; NIHR HPRUs in Chemical and Radiation Threats and Hazards, and Environmental Exposures and Health; the British Heart Foundation Centre for Research Excellence at Imperial College London (RE/18/4/34215); and the UK Dementia Research Institute at Imperial (MC_PC_17114). We thank The Huo Family Foundation for their support of our work on COVID-19.

We thank key collaborators on this work – Ipsos: Kelly Beaver, Sam Clemens, Gary Welch, Nicholas Gilby, Kelly Ward and Kevin Pickering; School of Public Health, Imperial College London: Graham Blakoe, Eric Johnson, Rob Elliott; Institute of Global Health Innovation at Imperial College: Gianluca Fontana, Sutha Satkunarajah, Didi Thompson and Lenny Naar; Molecular Diagnostic Unit, Imperial College London: Prof. Graham Taylor; North West London Pathology and Public Health England for help in calibration of the laboratory analyses; Patient Experience Research Centre at Imperial College and the REACT Public Advisory Panel; NHS Digital for access to the NHS register; and the Department of Health and Social Care for logistic support. SR acknowledges helpful discussion with attendees of meetings of the UK Government Office for Science (GO-Science) Scientific Pandemic Influenza – Modelling (SPI-M) committee.

## Supplementary Tables and Figures

**Supplementary Table 1.** The unweighted and weighted prevalence of swab-positivity across rounds 5 to 10 of REACT-1.

* Rounds 1 to 4 were conducted between 1/5/2020 and 8/9/2020 and found 473 positives from 596,565 tested swabs.

** Includes a small number of samples from previous days.

Supplementary Table 1 is available in this <u>Spreadsheet</u>.

**Supplementary Table 2.** The unweighted and weighted prevalence of swab-positivity by socio-demographic variables for rounds 5 to 10.

* We present weighted prevalence if the number of positives in a category is 10 or more.

Supplementary Table 2 is available in this <u>Spreadsheet</u>.

**Supplementary Table 3.** Odds ratios for rounds 5 to 10, and an average weighted inversely to variance for core variables. Models were adjusted for: 1) sex and age; 2) mutually adjusted for sex, age group, region, key worker status, ethnicity, household size, and deprivation index. The deprivation index is based on the Index of Multiple Deprivation (2019) at the lower super output area.

* HCW/CHW: health care worker/care home worker; Not FT, PT SE: not self-employed, full-time, part-time, self-employed.

Supplementary Table 3 is available in this <u>Spreadsheet</u>.

**Supplementary Table 4.** The unweighted and weighted prevalence of swab-positivity for detailed ethnicity subgroups for rounds 5 to 10.

* We present weighted prevalence if the number of positives in a category is 10 or more.

* Supplementary Table 4 is available in this <u>Spreadsheet</u>.

**Supplementary Table 5.** Odds ratios for rounds 5 to 10, and an average weighted inversely to variance for detailed subgroups of ethnicity. Models were adjusted for: 1) sex and age; 2) mutually adjusted for sex, age group, region, key worker status, and deprivation index. The deprivation index is based on the Index of Multiple Deprivation (2019) at the lower super output area.

* HCW/CHW: health care worker/care home worker; Not FT, PT SE: not self-employed, full-time, part-time, self-employed.

Supplementary Table 5 is available in this <u>Spreadsheet</u>.

**Supplementary Table 6.** The unweighted and weighted prevalence of swab-positivity for detailed categories of occupation for round 5.

* We present weighted prevalence if the number of positives in a category is 10 or more.

** We keep the variable names the same as the question names in questionnaires.

Supplementary Table 6 is available in this <u>Spreadsheet</u>.

**Supplementary Table 7.** Odds ratios for rounds 5 to 10, and an average weighted inversely to variance for detailed subgroups of occupations. Models were adjusted for: 1) sex and age; 2) mutually adjusted for sex, age group, region, ethnicity, and deprivation index. The deprivation index is based on the Index of Multiple Deprivation (2019) at the lower super output area.

Supplementary Table 7 is available in this <u>Spreadsheet</u>.

**Supplementary Table 8.** Odds ratios for rounds 9 and 10, and an average weighted inversely to variance for linked vaccination data. Models were: 1) mutually adjusted for sex, age group, region, key worker status, ethnicity, and deprivation index; 2) mutually adjusted for sex, age group, region, key worker status, ethnicity, deprivation index and vaccination status. The deprivation index is based on the Index of Multiple Deprivation (2019) at the lower super output area.

* HCW/CHW: health care worker/care home worker; Not FT, PT SE: not self-employed, full-time, part-time, self-employed.

**The sample size for linked vaccination data is smaller than REACT data used in previous regression analysis. The sample size for round 9 is 114,747, and for round 10 is 98,983.

Supplementary Table 8 is available in this <u>Spreadsheet</u>.

**Supplementary Figure 1.**
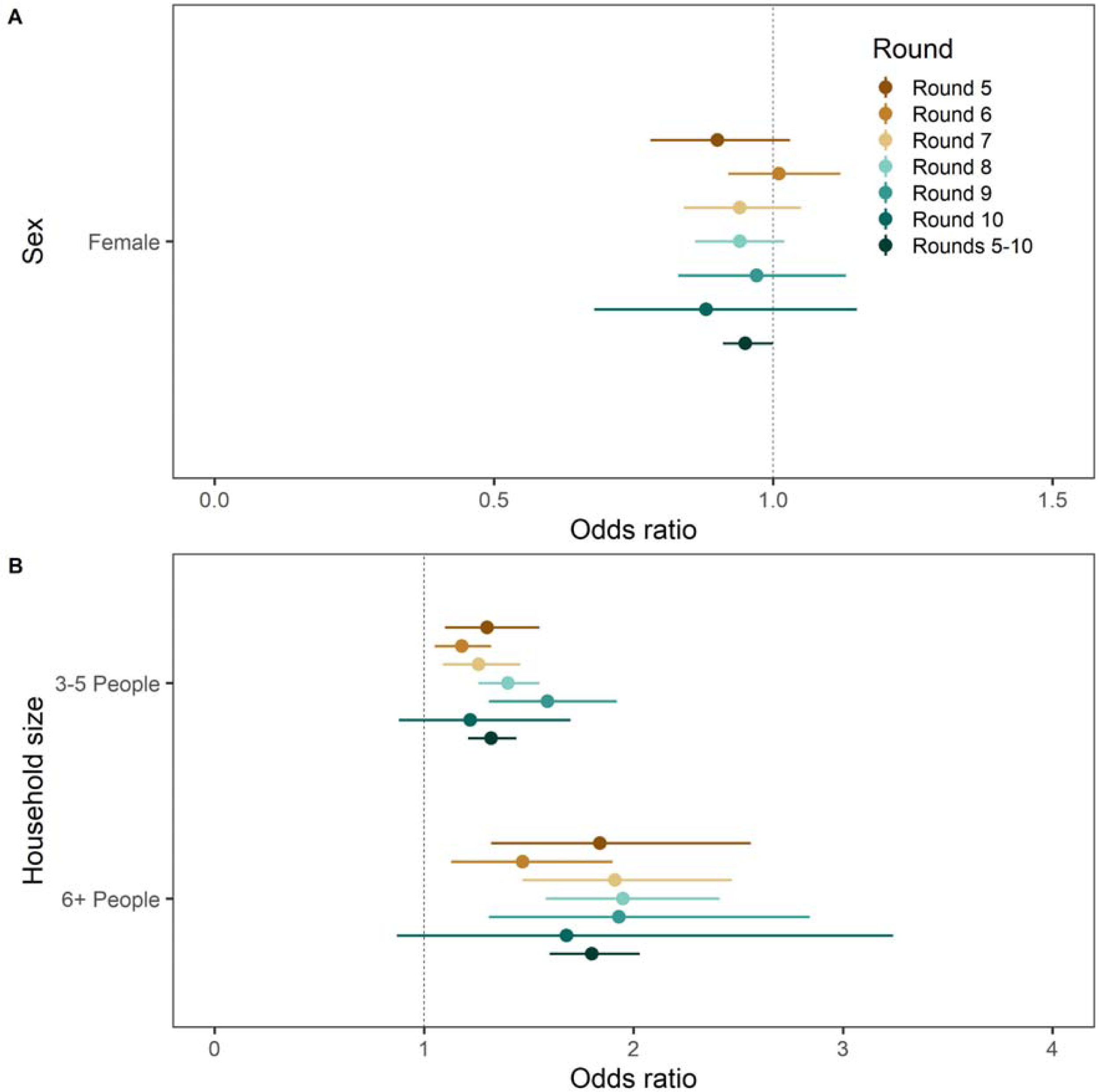
**A.** Forest plot of sex for rounds 5 to 10 and an average across rounds weighted inverse to variance (rounds 5 to 10). The reference group is not shown in the figure and is male. **B.** Forest plot of household size for rounds 5 to 10 and an average weighted across rounds inverse to variance (rounds 5 to 10). The reference group is not shown in the figure and is households with 1-2 people. Individual round odds ratios are mutually adjusted for sex, key worker status, broad categories of ethnicity, household size and neighbourhood deprivation.

