## Supplementary figures and images for "Health inequalities in SARS-CoV-2 infection during the second wave in England: REACT-1 study"

### Supplementary Figure 1

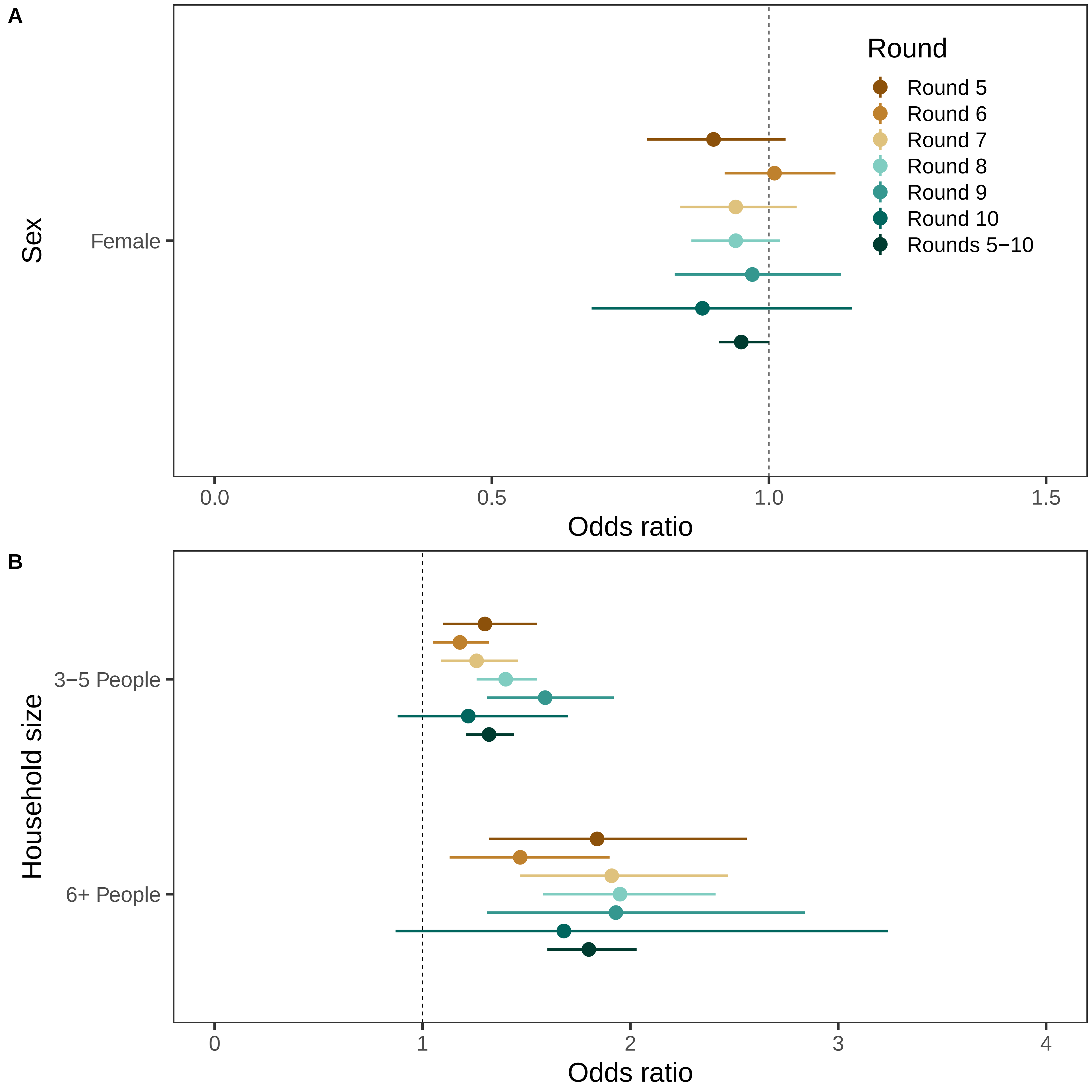
